# A triple bottom line approach to antimicrobial stewardship: the effects of simplifying treatment

**DOI:** 10.1101/2023.12.21.23300392

**Authors:** Michelle Balm, Olivia Bupha-Intr, Tanya Sinha, Matthew Kelly, Lucy Stewart, Ruth Stephen, Tim Blackmore, Max Bloomfield

## Abstract

**Aim:** Our antimicrobial guidelines (AGs) were changed in 2021 to recommend once-daily ceftriaxone in place of three-times-daily cefuroxime as preferred cephalosporin. This analysis sought to assess the effects of this on incidence of *Clostridioides difficile* infection (CDI), third-generation cephalosporin resistant Enterobacterales (3GCR-E), and resource utilisation.

**Method:** Before and after analysis of 30-day CDI and 3GCR-E incidence following receipt of cefuroxime/ceftriaxone pre- and post-AG change. Total nursing time and waste production relating to cefuroxime/ceftriaxone delivery were calculated pre- and post-change.

**Results:** CDI incidence was 0.6% pre- and 1.0% post-change (adjusted odds ratio [aOR] 1.44, p=0.07) and 3GCR-E incidence 3.5% and 3.1% (aOR 0.90, p=0.33). Mean per-quarter estimated nursing administration time decreased from 2065 to 1163 hours (902 nurse-hour reduction) and antibiotic-related waste generation from 1131kg to 748kg (383kg reduction). Overall days of therapy per-quarter of cefuroxime/ceftriaxone were unchanged between periods.

**Conclusion:** This simplification of our AG from a three-times-daily to a once-daily antibiotic resulted in considerable savings for our hospital (roughly 1.7 full-time equivalent nurses and over a tonne of waste yearly), with no significant increases in CDI or 3GCR-E. The impact of dosing schedules on non-antibiotic-spectrum factors, such nursing time and resource usage, is worthy of consideration when designing AGs.

## Introduction

A core tool in hospital antimicrobial stewardship (AMS) is the development of syndrome-specific antimicrobial guidelines (AGs).^1^ Guideline-adherent empirical therapy is associated with reduced treatment failure, length of stay (LOS) and mortality.^2^ Guideline development typically focuses on antimicrobial spectrum, route of administration, dose and duration. We believe that AMS should also promote responsible and sustainable use of limited resources including nursing time, patient comfort and convenience, and consumption of single-use plastic destined for landfill. This has parallels to the business concept of the ‘triple bottom line’, whereby success is measured not solely based on profit, but also in relation to the impact on people and the planet.

Using these principles, the AMS Committee at our institution changed the AGs in quarter 4 (Q4) 2021 to replace cefuroxime 1.5 g intravenous (IV) 8-hourly with ceftriaxone 2 g IV once daily as the preferred cephalosporin for empiric treatment of a number of indications. Ceftriaxone was already the recommended beta-lactam for moderate-severe community acquired pneumonia. This change generated considerable debate within the committee, particularly regarding the potential to drive antimicrobial resistance and *Clostridioides difficile* infection (CDI). Balancing this potential risk were the likely benefits to the hospital system in terms of simplicity (to promote adherence), nursing time saved and reduction in consumables. Convenience for patients was a major consideration with less disruption to rest and fewer missed or late doses. The aim of this report is to describe the positive and negative impacts of the change two years after implementation.

## Methods

### Setting

Wellington Regional Hospital (WRH) is a 484-bed acute care tertiary hospital which covers all specialties except plastic surgery and rheumatology, and is aligned with Kenepuru Hospital (KPH), a 131-bed community hospital. These hospitals service a population of around 500,000 people.

In Q4 2021, in addition to the AG changes described above, piperacillin-tazobactam was changed to cefepime for febrile neutropenia and severe hospital-acquired infection, and piperacillin-tazobactam changed to amoxicillin-clavulanate for mild-moderate hospital-acquired infection. There was widespread internal communication regarding these changes, and our AGs were updated on our hospital intranet and mobile app.^3^

### Data extraction and creation of cohorts

Medication dispensing at WRH/KPH uses BD Pyxis™, which permits data extraction for all individual dispensing events. The exception to this is the Emergency Department (ED), for which dispensing records were unavailable. The Pyxis data were used to create two cohorts for analysis: 1) patients aged >16 years who received at least 2 consecutive days of either IV cefuroxime or IV ceftriaxone; 2) patients aged >16 years who received at least 2 consecutive days of any other antibiotic via any route. Patients were excluded from this group if they had received cefuroxime, ceftriaxone, piperacillin-tazobactam, cefepime or amoxicillin-clavulanate within 30 days before or after the start of their course. The intention of the second cohort was to provide a comparator group with antibiotic use that was relatively unaffected by AG changes. Patient demographics and admission information were extracted from the hospital data warehouse, and microbiology data were extracted from the Awanui Laboratories Wellington laboratory information system, which provides both hospital and community microbiology services for the entire region.

### Definitions

The pre-change period was defined as 2019 Q1 to 2021 Q3, and the post-change period as 2022 Q1 to 2023 Q3. Q4 2021 was excluded because the new AGs were only partially embedded at this time. For each patient incident CDI was defined as either a positive direct fecal toxin enzyme immunoassay result or a positive toxigenic culture result within 2-30 days of the start of the antibiotic course. Incident ESBL/third-generation cephalosporin-resistant Enterobacterales (ESBL/3GCR-E) were defined as isolation of one of these organisms from any sample type within the same time window. An overnight dose of an antibiotic was defined as a dispensing event between 10pm and 6am.

### Estimates of administration time and waste

The time taken to administer a dose of IV antibiotics was estimated at 22 minutes.^4^ Weight estimates were obtained by collecting all the waste (antibiotic vials, giving sets, tubing, syringes, gloves, etc.) associated with delivery of each antibiotic over a 24-hour period and then dividing by the number of doses to generate an average weight per dose. Given the scarcity of patients on cefuroxime in the hospital, these weights were estimated from IV amoxicillin-clavulanate, which has the same dose frequency and almost exactly the same use of vials and consumables. This gave a weight of 84.5 g and 239.1 g for a 2 g dose of ceftriaxone via push and infusion, respectively, and 103.3 g and 195.6 g for cefuroxime. The infusion weight was used to estimate the overall waste generation because this is the predominant mode of delivery in our institution.

### Analysis

Outcomes of interest were: 1) the 30-day incidence of CDI and ESBL/3GCR-E in each cohort; 2) estimated total nursing time spent administering IV antibiotics, estimated total IV-associated waste generation, and total overnight doses administered in the cephalosporin cohort. These outcomes were compared across the time periods, with the Chi-squared test used to compare categorical variables, and the Mann-Whitney-U test for continuous variables. A multivariate logistic regression model was created to determine which covariates in the cephalosporin cohort were independently associated with increased odds of CDI and ESBL/3GCR-E. Analysis was performed in Stata 17 (College Station, Texas). This analysis was part of the routine ongoing monitoring activities undertaken at our hospital by the AMS committee, which is an ongoing Continuous Quality Improvement project to inform future guidelines. It was therefore out of scope for Health and Disability Ethics Committee review, as this analysis is classified as an ‘audit or related activity’. Hospital Clinical Audit Committee approval was gained.

## Results

The characteristics of the cephalosporin cohort are shown in Table 1. In the pre-change period, 85.7% of the cohort received cefuroxime, whereas in the post-change period 94.2% received ceftriaxone, consistent with the change in AGs. The pre- and post-change groups were otherwise broadly similar, other than a decrease in general surgical patients and an increase in general medical patients in the post-change period. Mean days of therapy (DOT) with cefuroxime/ceftriaxone within 30 days of the initial dose were similar across time periods (4.1 vs 4.0, p=0.01). CDI incidence was 0.6% in the pre-change, versus 1.0% in the post-change period (p=0.02). Isolation of ESBL/3GCR-E within 30-days was 3.5% pre-change versus 3.1% (p=0.31) post-change. Characteristics of the comparator group are shown in Table 2. The major difference compared to the cephalosporin cohort was that there were fewer general surgical patients (6.6% and 5.1% of the cohort pre- and post-change) and more subspecialty surgical patients (39.8% and 46.5%). The incidence of CDI was lower than the cephalosporin cohort and did not change between time periods (0.2% and 0.2%, p=0.68) and incidence of ESBL/3GCR-E was not significantly different between periods (1.4% versus 1.7%, p=0.13). Figure 1. shows CDI incidence over time in the two cohorts. The incidence increased prior to the AG change in the cephalosporin cohort, appeared to peak shortly after the change, and decreased after this, whereas the incidence appeared stable across both time periods in the comparator group.

**Table 1.** Characteristics and outcomes of the cephalosporin cohort pre- and post-change in antimicrobial guidelines.

|  | Pre-change |  | Post-change |  | p-value |
| --- | --- | --- | --- | --- | --- |
| Total patients | 8342 |  | 5107 |  |  |
| Received cefuroxime | 7150 | 85.7% | 296 | 5.8% |  |
| Received ceftriaxone | 1192 | 14.3% | 4811 | 94.2% |  |
| Age (years), median, IQR | 60.3 | (39.3,75.5) | 64.9 | (47.1,76.9) | <0.01 |
| Female | 4616 | 55.3% | 2675 | 52.5% | <0.01 |
| Ethnicity |  |  |  |  | 0.01 |
| Other or unknown | 1486 | 17.8% | 830 | 16.3% |  |
| NZ Māori | 1099 | 13.2% | 679 | 13.3% |  |
| Pacific Island | 778 | 9.3% | 569 | 11.1% |  |
| NZ European | 4335 | 52.0% | 2643 | 51.8% |  |
| Asian | 644 | 7.7% | 386 | 7.6% |  |
| Specialty grouped |  |  |  |  | <0.01 |
| General medicine | 2273 | 27.2% | 1700 | 33.3% |  |
| Subspecialty medicine | 429 | 5.1% | 330 | 6.5% |  |
| Hematology/Oncology | 669 | 8.0% | 471 | 9.2% |  |
| Intensive care | 181 | 2.2% | 191 | 3.7% |  |
| General surgery | 2591 | 31.1% | 1270 | 24.9% |  |
| Subspecialty surgery | 891 | 10.7% | 596 | 11.7% |  |
| Older persons' health | 70 | 0.8% | 42 | 0.8% |  |
| Women's health | 1023 | 12.3% | 383 | 7.5% |  |
| Emergency department<sup>a</sup> | 202 | 2.4% | 119 | 2.3% |  |
| Other | 13 | 0.2% | 4 | 0.1% |  |
| Hospitalized in the last 365 days | 4139 | 49.6% | 2588 | 50.7% | 0.21 |
| CDI in prior 365 days | 39 | 0.5% | 43 | 0.8% | <0.01 |
| Cefuroxime/ceftriaxone DOT within 30 days | 4.1 | 2.9 | 4.0 | 2.7 | 0.01 |
| Incident CDI within 30 days | 54.0 | 0.6% | 52 | 1.0% | 0.02 |
| ESBL/3GCR-E culture within 30 days | 289 | 3.5% | 159 | 3.1% | 0.31 |

IQR, inter quartile range; CDI, *Clostridioides difficile* infection; DOT, days of therapy; ESBL/3GCRE-E, extended-spectrum beta-lactamase or third generation cephalosporin-resistant Enterobacterales.
<sup>a</sup>Although emergency department Pyxis data were unavailable, some patients were still coded as being under emergency once they had been admitted to a ward and received their first antibiotic dose.

**Table 2.**
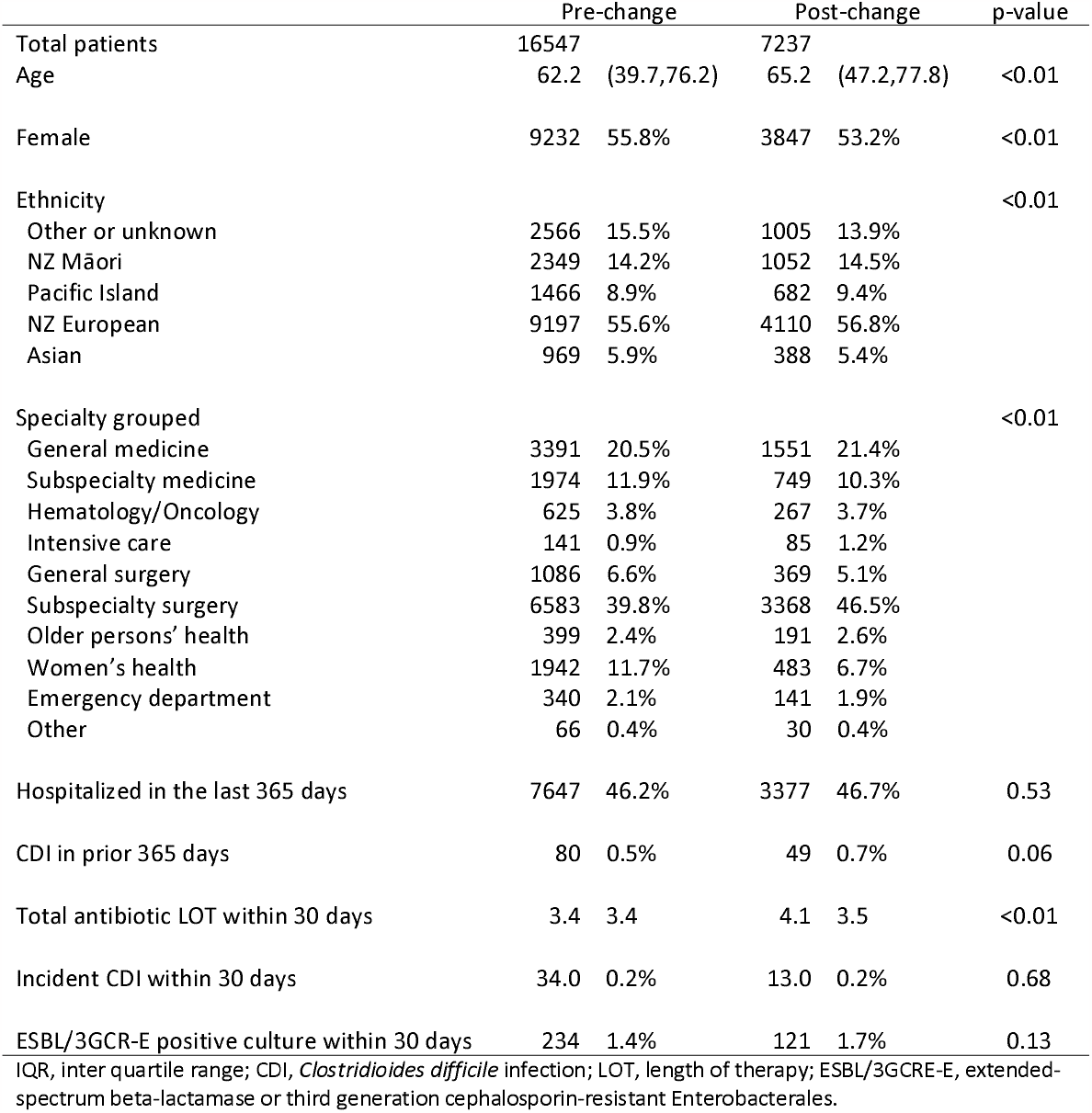
Characteristics and outcomes of the comparator cohort pre- and post-change in antimicrobial guidelines.

|  | Pre-change |  | Post-change |  | p-value |
| --- | --- | --- | --- | --- | --- |
| Total patients | 16547 |  | 7237 |  |  |
| Age | 62.2 | (39.7,76.2) | 65.2 | (47.2,77.8) | <0.01 |
| Female | 9232 | 55.8% | 3847 | 53.2% | <0.01 |
| Ethnicity |  |  |  |  | <0.01 |
| Other or unknown | 2566 | 15.5% | 1005 | 13.9% |  |
| NZ Māori | 2349 | 14.2% | 1052 | 14.5% |  |
| Pacific Island | 1466 | 8.9% | 682 | 9.4% |  |
| NZ European | 9197 | 55.6% | 4110 | 56.8% |  |
| Asian | 969 | 5.9% | 388 | 5.4% |  |
| Specialty grouped |  |  |  |  | <0.01 |
| General medicine | 3391 | 20.5% | 1551 | 21.4% |  |
| Subspecialty medicine | 1974 | 11.9% | 749 | 10.3% |  |
| Hematology/Oncology | 625 | 3.8% | 267 | 3.7% |  |
| Intensive care | 141 | 0.9% | 85 | 1.2% |  |
| General surgery | 1086 | 6.6% | 369 | 5.1% |  |
| Subspecialty surgery | 6583 | 39.8% | 3368 | 46.5% |  |
| Older persons' health | 399 | 2.4% | 191 | 2.6% |  |
| Women's health | 1942 | 11.7% | 483 | 6.7% |  |
| Emergency department | 340 | 2.1% | 141 | 1.9% |  |
| Other | 66 | 0.4% | 30 | 0.4% |  |
| Hospitalized in the last 365 days | 7647 | 46.2% | 3377 | 46.7% | 0.53 |
| CDI in prior 365 days | 80 | 0.5% | 49 | 0.7% | 0.06 |
| Total antibiotic LOT within 30 days | 3.4 | 3.4 | 4.1 | 3.5 | <0.01 |
| Incident CDI within 30 days | 34.0 | 0.2% | 13.0 | 0.2% | 0.68 |
| ESBL/3GCR-E positive culture within 30 days | 234 | 1.4% | 121 | 1.7% | 0.13 |

IQR, inter quartile range; CDI, *Clostridioides difficile* infection; LOT, length of therapy; ESBL/3GCRE-E, extended-spectrum beta-lactamase or third generation cephalosporin-resistant Enterobacterales.

**Figure 1.**
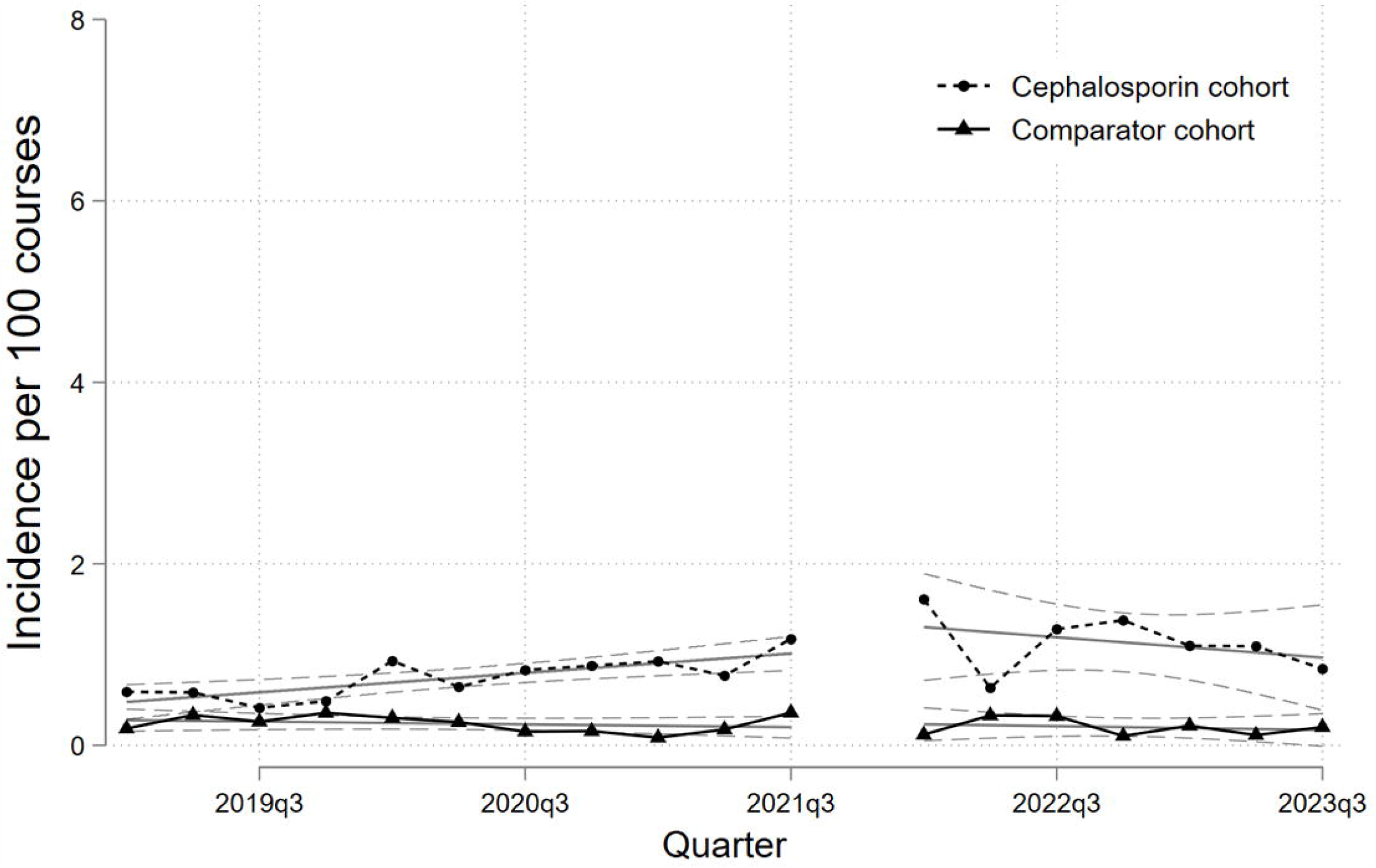
Thirty-day incidence of *Clostridioides difficile* infection by quarter in the cephalosporin and comparator cohorts, with linear trend line and 95% confidence interval.

Table 3. shows the multivariate analysis results for CDI. Several covariates were associated with increased odds of CDI in the cephalosporin cohort, with the strongest being a prior diagnosis of CDI within the last year (aOR 11.3, 95%-CI 5.25-24.33, p<0.01), and cefuroxime/ceftriaxone DOT within 30 days of index dose (aOR 1.07 for each additional DOT, 95%-CI 1.02-1.13, p=0.01). The odds of CDI in the post-change period was no longer statistically significantly higher than the pre-change period (aOR 1.44, 95%-CI 0.97-2.12, p=0.07). The results of the multivariate analysis for ESBL/3GCR-E showed an aOR of 0.91 (95%-CI 0.74-1.11, p=0.34) for incident ESBL/3GCR-E in the post-change period (Supplementary materials).

**Table 3.** >Multiple logistic regression model for odds of incident CDI in the cephalosporin cohort, by different patient characteristics.

|  | aOR | 95% CI | p-value |
| --- | --- | --- | --- |
| Age | 1.02<sup>a</sup> | (1.00, 1.03) | 0.01 |
| Female | 1.68 | (1.12, 2.50) | 0.01 |
| Ethnicity |  |  |  |
| Other or unknown | 1.00 |  |  |
| NZ Māori | 1.40 | (0.71, 2.76) | 0.34 |
| Pacific Island | 0.32 | (0.09, 1.09) | 0.07 |
| NZ European | 1.04 | (0.61, 1.77) | 0.89 |
| Asian | 0.64 | (0.21, 1.90) | 0.42 |
| Specialty grouped |  |  |  |
| General medicine | 1.00 |  |  |
| Subspecialty medicine | 1.74 | (0.76, 3.96) | 0.19 |
| Hematology/Oncology | 2.19 | (1.15, 4.16) | 0.02 |
| Intensive care | 2.52 | (0.94, 6.77) | 0.07 |
| General surgery | 1.12 | (0.62, 2.01) | 0.72 |
| Subspecialty surgery | 2.40 | (1.32, 4.37) | <0.01 |
| Older persons' health | 2.68 | (0.77, 9.25) | 0.12 |
| Women's health | 0.41 | (0.09, 1.90) | 0.26 |
| Emergency department | -<sup>b</sup> |  |  |
| Other | -<sup>b</sup> |  |  |
| Hospitalized in the last 365 days | 1.41 | (0.92, 2.15) | 0.12 |
| CDI in prior 365 days | 11.30 | (5.25, 24.33) | <0.01 |
| Cefuroxime/ceftriaxone DOT within 30 days | 1.07<sup>a</sup> | (1.02, 1.13) | 0.01 |
| Post-change period patient | 1.44 | (0.97, 2.12) | 0.07 |

aOR, adjusted odds ratio; CDI, *Clostridioides difficile* infection; DOT, days of therapy
<sup>a</sup>Odds ratio here represents increased odds per unit increase in the variable.
<sup>b</sup>Insufficient outcomes in these groups to generate an odds ratio.

Table 4. shows the hospital-wide effects of the change in cephalosporin AGs between time periods. The hospital was busier in the post-change period (additional 2487 occupied bed days per quarter), but despite this, total cefuroxime/ceftriaxone DOT was very similar (2595 DOT versus 2599 DOT per quarter). As expected, there was a marked decrease in mean cefuroxime DOT per quarter (2067 versus 105) and increase for ceftriaxone (528 versus 2494). There was a large decrease in mean combined cefuroxime/ceftriaxone dosing events per quarter (5633 to 3172, decrease of 2461), which translated to large reductions in estimated nursing time consumption (2065 hours per quarter versus 1163, reduction of 902) and estimated waste generated for infusion delivery (1131kg to 748kg, reduction of 383kg). The mean number of overnight dosing events also decreased substantially (1777 versus 588, decrease of 1189 per quarter).

**Table 4.**
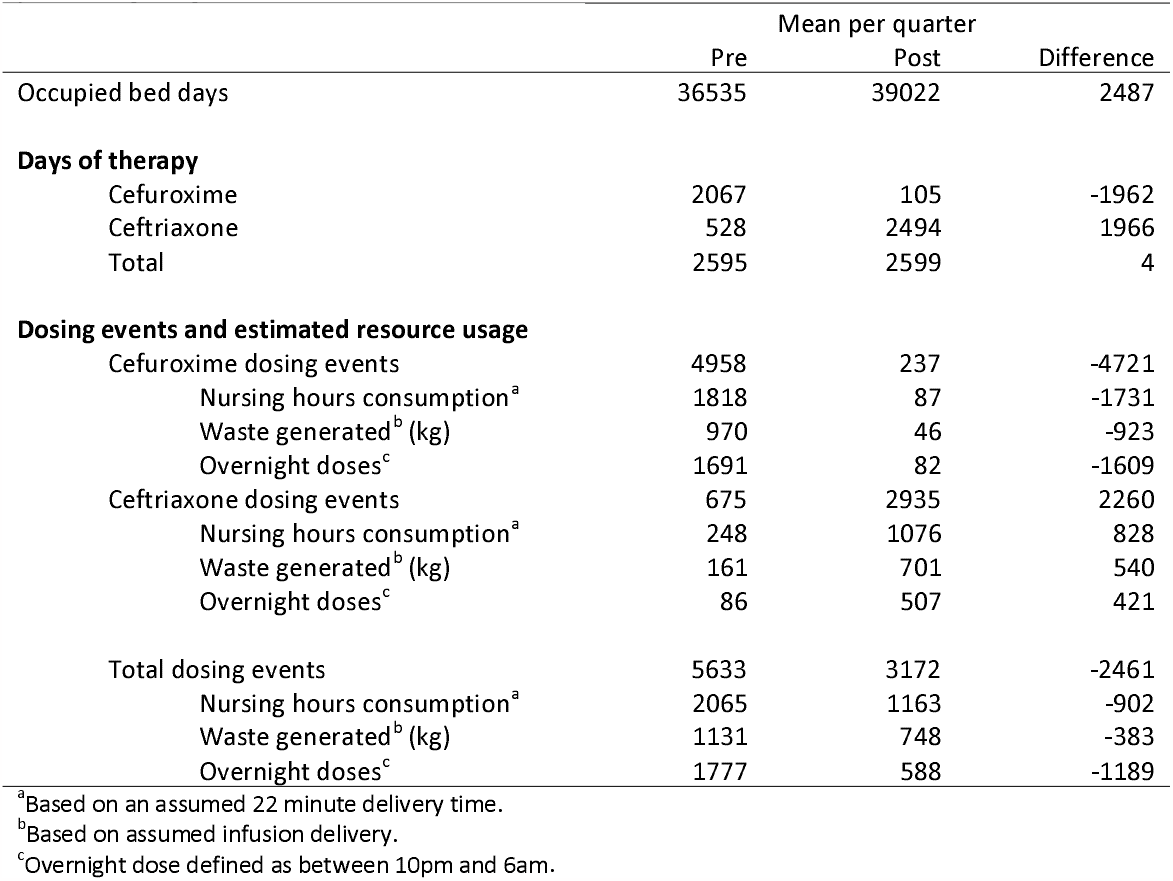
Mean usage and resource consumption for cefuroxime and ceftriaxone per quarter pre- and post-change in guidelines.

## Discussion

This study demonstrates widespread hospital acceptance of pragmatic guidelines which take into consideration nursing workloads and patient convenience, with no statistically significant increase in CDI or resistant organisms. We argue that these principles should be significant considerations for the development of AGs, and that AMS should consider this ‘triple bottom line’ framework, rather than focusing solely on the implications of antibiotic spectrum. Our programme emphases the importance of ongoing monitoring of resistant organisms and CDI, to ensure that we do not cause harm to the wider patient group in the name of nursing and individual patient convenience.

NZ has a health workforce crisis, and it is especially important that nursing time is used optimally with improvements in workflow implemented where possible.^5^ A major benefit with the guideline change was the reduction in nursing time spent preparing and delivering IV antibiotics, primarily due to recommending a once-daily antibiotic. A mean estimated reduction of 902 nursing hours per quarter was realised, which equates to almost 10 hours per day, or roughly 1.7 full-time equivalent. In a period when there is a shortage of nurses, the ability to reallocate scarce nursing resource to other care priorities is a considerable advantage. Furthermore, overstretched nurses are known to be less compliant with hand hygiene and other infection control principles, so a reduction in workload may be beneficial in this regard, which is in line with the goals of AMS.^6^

Both ceftriaxone 2 g and cefuroxime 1.5 g can be administered as either a push (3-5 minutes for cefuroxime, 4-8 minutes for ceftriaxone) or via a 30-minute infusion.^7^ While push administration is associated with less waste and overall time of delivery, observation during this study showed that infusion remains standard practise in our hospital. In several studies, an IV push antibiotic was associated with reduced time spent delivering doses compared to infusions, faster time to first dose and reduced consumables and costs per dose event.^8–10^ Antibiotic infusion may also result in wastage of up to 24% of the dose if the residual volume is not flushed, an issue not present for bolus dosing.^10^ A program to encourage IV push of appropriate antibiotics may be a useful next step at our institution, although the convenience of being able to perform other tasks once an infusion has been hung is a common advantage cited by our nurses, so the reported time savings of push administration may be less relevant, although would still result in savings in waste.

Our findings suggest that a significant reduction in waste can be achieved with once-daily antibiotics such as ceftriaxone. We estimated that up to 383kg per quarter of consumable waste, or over a tonne of waste per year, was saved by the guideline changes (assuming infusion delivery; the quarterly estimate would be 297kg if all delivery was via push). Hospital waste contributes almost 5% of global greenhouse gas emissions and uses substantial plastic consumables.^11,12^ In NZ, most of the consumables are imported, adding to the emission footprint. Minimising single-use consumables (IV tubing, syringes, flush vials, plastic fluid bags) should be a priority for hospitals given the threats to human health from environmental contamination and climate change.^11,12^ It should be noted that the time savings and waste generated are based on per-dose estimates and the final numbers should not be considered exact, however they are likely to be a good indication of the magnitude of potential savings.

Use of a once-a-day antibiotic improves patient comfort and convenience, and should eliminate the need to give overnight doses, other than the first dose in patients presenting overnight. There was a reduction of 1189 doses given overnight compared to the pre-change period. However, 22.6% of doses were still given in the overnight period. This suggests a failure to chart antimicrobials at more nurse- and patient-friendly times by doctors, who may not think of the negative impact on patient sleep and workload for nurses overnight – a time where staffing levels are lower. Interventions aimed at changing workflows including time of medication administration have been shown to be successful at improving sleep disruption for patients.^13,14^ This is an area to target for prescriber education in our institution. A further advantage of once daily dosing is that it frees up patient time for other important aspects of their care, such as allied health, radiology, or other therapies.

Critics of our approach may emphasise the potential harms of ceftriaxone when compared to cefuroxime due to its broader spectrum. Although there are theoretical concerns regarding the emergence of antimicrobial resistance and CDI with use of broad-spectrum antibiotics, studies are heterogenous and do not provide simple answers. Antibiotics clearly have a profound effect on the microbiome, and this can be persistent but the effect is complex.^15^ Cephalosporins are associated with increased carriage of AmpC and ESBL-producing organisms.^16^ Ceftriaxone and cefuroxime both promote increases in *bla*_*CTX-M*_ abundance, with one study demonstrating a 22% per day increase in CTX-M genes in patients exposed to cefuroxime and 10% increase for patients on ceftriaxone.^17^ Short course antibiotics impact less on AMR genes in gut microbiome, and some studies have shown that the spectrum of antibiotics has less impact than duration of exposure, thus the impact on the microbiome depends on a range of factors including duration, route of administration as well as antibiotic spectrum.^15,18,19^ Our data to date did not show an increase in ESBL-producing organisms relating to the change in cephalosporin AGs.

Data on antibiotic-specific risk for CDI is also challenging due to heterogenous methods and study design. Miller recently showed that cefuroxime has a similar odds ratio for CDI to third generation cephalosporins, however, their study used cefpodoxime rather than ceftriaxone.^20^ The EUCLID study showed CDI risk was based on complex factors, not just antibiotic selection, and were unable to show a clear association with overall cephalosporin prescribing.^21^ An older systematic review and meta-analysis showed an OR of 3.2 for ceftriaxone compared to 2.23 for cefuroxime for CDI.^22^ Our data showed a non-statistically significant increase in 30-day CDI incidence in the post-change period. However, the incidence appeared to have been gradually increasing over time prior to the change, with a peak shortly after the change, and subsequent return towards baseline levels. Ongoing surveillance will provide more robust data regarding the effects of guideline changes but so far there are no convincing clinical data that establishes ceftriaxone to be a higher risk antibiotic than cefuroxime for the risk of CDI or antimicrobial resistance.

The overall impact of initial empirical antimicrobials used in a hospital setting on the development of resistant gram negatives and CDI is likely to be less significant than the ongoing antimicrobial use in terms of spectrum and duration, and that of community prescribing, which in NZ accounts for around 95% of human antibiotic consumption.^23^ In an AMS programme that prioritizes early review, IV to oral switch, and short courses, we have shown that taking a pragmatic initial approach helps our nursing colleagues, patients and the environment. Whilst it is clear that antibiotics are a precious and limited resource, we believe it is important to consider the other resources that antibiotic administration requires. Analysis of antibiotic use should be broadened to include a ‘triple bottom line’ approach, considering impact on nursing workload, patient comfort, and environmental impact. We believe that these are important considerations for institutional guidelines and should form part of any contemporary AMS strategy. Integral to these concepts is actively monitoring the positive and negative impact of changes and responding to the findings objectively.

## Data Availability

All data produced in the present work are contained in the manuscript.

## Competing interests

The authors have no conflicts of interest to declare.

## Funding

This work was supported by internal departmental funds.

## Acknowledgements

The authors would like to thank Matthew Ordish for assistance in accessing Pyxis data and Peter Wash for assistance in accessing hospital bed occupancy and demographic data.

## Notes

### Competing Interest Statement

The authors have declared no competing interest.

### Funding Statement

This study did not receive any funding.

### Author Declarations

The Health and Disability Ethics Committees (HDECs) of New Zealand waived ethical approval for this work.

