## Supplemental Table 1 for "A triple bottom line approach to antimicrobial stewardship: the effects of simplifying treatment"

| Supplementary Table 1. Multiple logistic regression model for odds of ESBL/3GCR-E in the cephalosporin cohort, by different patient characteristics | | | |
| --- | --- | --- | --- |
|  | Coefficient | 95% CI | p-value |
| Age | 1.02^a^ | (1.01, 1.02) | 0.00 |
| Female | 1.24 | (1.02, 1.50) | 0.03 |
| Ethnicity |  |  |  |
| Other or unknown | 1.00 |  |  |
| NZ Maori | 1.00 | (0.69, 1.44) | 0.98 |
| Pacific Island | 0.86 | (0.57, 1.30) | 0.47 |
| NZ European | 0.99 | (0.76, 1.29) | 0.92 |
| Asian | 1.64 | (1.11, 2.41) | 0.01 |
| Specialty grouped |  |  |  |
| General medicine | 1.00 |  |  |
| Subspecialty medicine | 1.00 | (0.62, 1.61) | 1.00 |
| Haematology/Oncology | 1.46 | (1.03, 2.08) | 0.04 |
| Intensive care | 2.22 | (1.36, 3.62) | 0.00 |
| General surgery | 1.41 | (1.09, 1.83) | 0.01 |
| Subspecialty surgery | 1.42 | (1.03, 1.97) | 0.03 |
| Older persons’ health | 0.96 | (0.37, 2.48) | 0.93 |
| Women’s health | 0.64 | (0.35, 1.15) | 0.14 |
| Emergency department^b^ | 0.69 | (0.30, 1.61) | 0.39 |
| Other | 1.00^b^ |  |  |
| Hospitalised in the last 365 days | 1.04 | (1.01, 1.08) | 0.02 |
| Cefuroxime/ceftriaxone DOT within 30 days | 1.14^a^ | (1.12, 1.16) | 0.00 |
| Post-change period patient | 0.91 | (0.74, 1.11) | 0.34 |
| ESBL/3GCR-E, extended-spectrum beta-lactamase-producing or third generation cephalosporin-resistant Enterobacterales; aOR, adjusted odds ratio; DOT, days of therapy  ^a^Odds ratio here represents increased odds per unit increase in the variable.  ^b^Insufficient outcomes in these groups to generate an odds ratio. | | | |
